# Moving from theory to practice: exploring power and power sharing in participatory health research partnerships: a scoping review protocol

**DOI:** 10.1101/2022.11.21.22282508

**Authors:** Nino Burduladze, Laundette P. Jones, Brian D. Jones, Uchizi Msowoya, Jon Salsberg, Anna Whitney, Meghan Gilfoyle

**Author notes:** **Corresponding Author** (NB). These authors contributed equally to this work (Nino Burduladze, Laundette P. Jones, Brian D. Jones, Jon Salsberg, Meghan Gilfoyle). These authors also contributed equally to this work (Uchizi Msowoya, Anna Whitney).

## Abstract

**Introduction:** Participatory health research (PHR) as a research paradigm, guides the research process and strives to achieve positive change in society in the interest of people’s health. In this scoping review, PHR will be used as an umbrella term considering a wide range of collaborative research approaches in the health context. PHR is conducted ‘with’ or ‘by’ those it intends to benefit, as opposed to ‘on’ and ‘for’ them. Their involvement throughout the research process seeks to shift power and decision-making from where they traditionally lay within academia toward community, patient and public end-users. Research cannot be truly participatory without concurrently addressing power dynamics within the partnership and power imbalances in decision making. Therefore, power sharing can be defined as a major factor in building effective academic-community collaborations. This scoping review aims to identify, clarify, and map existing literature on power and power sharing in PHR from both theoretical and practical perspectives. Specifically, we will explore how is power conceptualised throughout the literature, and how power and power sharing are applied and addressed in real-life PHR partnerships.

**Materials and methods:** This scoping review will be conducted in accordance with the guidelines outlined in the Joanna Briggs Institute (JBI) Reviewer’s Manual. This scoping review will consider both empirical and non-empirical research that report on understanding power and power sharing in participatory health research partnerships. All appropriate studies will be retrieved from the following five electronic databases: PubMed, Scopus, Embase, PsycINFO, SocIndex. This review will be limited to articles published in English and from January 1998 to April 2022. As the scoping review aims to capture more than peer-reviewed and published literature, it will also include grey literature such as theses and dissertations, reports, conference proceedings, and editorials. Data from the included literature will be extracted based on the data extraction tool, defined in advance.

**Ethics and dissemination:** As primary data will not be collected, ethical approval is not required to conduct the scoping review. The findings of this study will be disseminated through peer-reviewed publications.

## Introduction

### Background

Participatory Health Research (PHR) is a research paradigm that aims to generate new knowledge leading to action for addressing social disparities in health outcomes (1). It can be seen as an umbrella term that encompasses a range of research approaches (2, 3), such as, integrated knowledge translation (IKT), a participatory approach that primarily partners with clinicians, service managers and policy makers, as well as patients and other end-users (4); public and patient involvement which focuses predominantly on the involvement of patients and their carers (5); and community-based participatory research (CBPR) that typically focuses on the involvement of communities and community-based stakeholders to address a wide range of problems ranging from individual care up to public health policy intervention (6). Regardless of the term used, all share a philosophy of inclusivity and end-user involvement throughout the research process ensuring that those who the research intends to benefit are at the heart of the research decision-making process (7).

The defining principle of PHR is to maximise the involvement of those whose life or work is the subject of research in all stages of the research process. PHR strives to harness equitable opportunity for all research partners (i.e., those with both experiential and methodological expertise) to be meaningfully involved in the research process to the greatest extent possible(8), while intentionally creating collaborative spaces (9) between those who typically have power and those that often do not (i.e., doctors together with patients, policy makers together with refugees, or academic researchers with community partners). Participatory processes help ensure ‘transparent involvement of different stakeholders’ (10) and that all voices are heard. Thus, through these processes, PHR enables a shift in power and decision-making from where they traditionally lay within academia towards community, patient and public end-users (11).

Achieving this level of participation at all stages of health research is challenging and affected by social hierarchies, gender, power, privilege, marginalisation and other factors(12). Power imbalances among academic researchers and non-academic participants, jeopardises the ability of PHR to bring about positive and long-term change (13). Research can entail different degrees of participation, but formal research involvement that does not concurrently address power dynamics among partners and power imbalances in decision making is not truly participatory (14). Thus, existing literature defines power sharing as a major factor in building effective academic-community collaborations (14). Recognising the importance of power and power sharing, a group of researchers from the International Collaboration for Participatory Health Research (ICPHR) created online workshops to explore how PHR projects, from a variety of global contexts, understand the dimensions of power and power sharing (13). Results from this qualitative study demonstrated that while participatory researchers acknowledge the importance of unpacking “power”, the concept is rarely discussed and addressed explicitly in research partnerships (13). Thus, although commonly discussed in the literature as an imperative part of the PHR process, gaps in understanding the practical aspects of power and power sharing persist, both conceptually and operationally. (11) (3) (10) (13).

### Objectives of scoping review

This scoping review aims to identify, clarify, and map existing literature on power and power sharing in PHR from both theoretical and practical perspectives. Specifically, we will explore how power is conceptualised throughout the literature, and how power and power sharing are operationalised in real-life participatory health research partnerships. In doing so, we will answer the following research question:

*“How is power conceptualised, operationalised and measured both theoretically and practically in participatory health research partnerships?”*

Accordingly, objectives of this scoping review are to:

1. Explore how power is conceptualised in PHR.
2. Understand how is power operationalised and measured in PHR.
3. Determine whether existing literature discusses power in PHR and if so, at what stage? (i.e., who holds funds, decisions about methodology, who gets to be co-investigator, etc.).
4. Report on how (if at all) authors suggest addressing power and power inequities in PHR partnerships and the barriers and facilitators of this.
5. Explore if there are any contextual differences to the above objectives (i.e., geographical, cultural, socioeconomic, demographics (e.g., race/ethnicity, etc.).

## Materials and methods

A scoping review was deemed appropriate for this research as it is considered as an effective tool for exploring, mapping and summarising the breadth of literature in a given field, when the scope of the review and research question are broad in focus (15). This scoping review will be conducted in compliance with the most up-to-date scoping review guidelines outlined in the Joanna Briggs Institute (JBI) Reviewer’s Manual (16). The JBI Reviewer’s Manual is premised on the guidelines established by Levac *et al*, (17), which is a further refined methodological framework from Arksey and O’Malley (18). We will incorporate the Population (or participants), Concept and Context (PCC) framework (16) to identify these elements of interest for this scoping review.

### Inclusion criteria

Detailed justification for inclusion and exclusion criteria is outlined below.

### Types of Participants

Only people over the age of 18 will be considered for the study’s sample. This is because there are different ethical considerations when involving those under the age of 18 in research (19). The partnership may include individuals or groups of people who are participating in health research and, directly or indirectly, are affected by the results and findings of the research.

### Concept

The main concept under investigation is power and power sharing, which will be examined in four ways. First, we will explore how literature conceptualises power, and whether different types or levels of power exist (e.g., power within, power with, power to, power over)(21, 22). The second aspect is surrounding how power is operationalised and measured among partners from both theoretical and practical perspectives, how partners perceive it, who oversees facilitating the process in a real-life setting, and how participants are engaged and involved in the PHR process. When we say theoretically, we mean identifying how authors would define the concept of power and power sharing as well its effects; while practically refers to how these theoretical insights are applied to sustain functioning research partnerships (23). Third, we will explore if power inequities are discussed in the literature and if/how authors sought to address these inequities. Finally, we will explore how all the above differ across varying contexts (i.e., geographical, cultural, socioeconomic etc.).

### Context

The context of interest for this scoping review is PHR acting as an umbrella term for other types of health-related collaborative research approaches, in which all aspects of the abovementioned concept will be examined. Further, any other forms of participatory research outside of the health context will be excluded.

This review will be limited to articles published from January 1998 until April 2022. This timeframe reflects the 1998 adoption of the North American Primary Care Research Group’s (NAPCRG) Policy Statement on *Responsible Research with Communities: Participatory Research in Primary Care* (24) after which the use of PHR principles in health-related research became more common (24).

As the reviewers only speak English, studies in languages other than English will be excluded. Furthermore, the scoping review’s context is PHR globally and internationally, thus, no country or geographical location restrictions will be used during the screening process.

### Types of sources

This scoping review will consider all types of quantitative, qualitative, and mixed-methods studies and reviews that report on understanding power and/or power sharing in participatory health research partnerships, respectively, responding to the review objectives outlined above.

Grey literature such as theses and dissertation, unpublished articles, opinion pieces, reports, conference proceedings, would also be considered. Besides, systematic reviews that meet the inclusion criteria will also be considered.

### Search strategy

All aspects of the search strategy development, including choosing the appropriate databases, will be carried out in conjunction with a faculty librarian from the University of Maryland, Baltimore.

To identify relevant studies and articles, five electronic databases will be searched, including PubMed, Scopus, Embase, PsycINFO, SocIndex. In addition, grey literature will be searched in Google Scholar and Open Grey databases.

Following the guidance of JBI (25), a three-step search strategy will be used. First, a preliminary search of PubMed, and Scopus will be conducted. This initial search will then be followed by an analysis of the text contained in the title, abstracts and index terms of retrieved papers. A second search using all identified keywords and index terms will be applied across all databases, including PubMed, Scopus, Embase, PsycINFO, SocIndex. Finally, we will review additional sources from the reference lists of the literature selected for full-text screening. The complete search strategy from the PubMed database can be found in Appendix I.

### Study selection

Following the search, results from all databases will be exported to Covidence software (26) for screening. Before each source is screened for eligibility, duplicates will be removed. The screening process for study selection will be conducted by two independent reviewers in two stages. First, two independent reviewers will screen the literature by titles and abstracts based on the predetermined inclusion criteria. Following, all included articles from the title and abstract screening will be screened at full text, documenting at this stage, the reasons for the exclusion. If the two independent screeners disagree, the final decision will be made by a third reviewer/arbitrator. At each review stage, all reviewers (including arbitrator) will select approximately 10% of the literature to review. They will then meet with the entire research team to discuss their thought process, decision-making, and revisit the eligibility criteria. Reviewers will then repeat this step, and will only proceed independently once an agreement rate of 75% or greater is achieved (16).

### Data extraction

Two independent reviewers will extract data from the included literature using the data extraction tool based on Peters at al.,(16) and refined by the reviewers. Extracted information will include, for example, publication details, general study details, and key findings relevant to the review objectives. See Table 2 below for the preliminary charting table. If there are any changes required during the data extraction process (i.e., key findings related to the concept of power, additional study details etc.), a detailed description of such changes will be reported in the full scoping review. Preferred Reporting Items for Systematic Reviews and Meta-Analyses flow chart (PRISMA) (found in Appendix II) (27) will be also used to identify the number of included and excluded articles at each stage of the review process.

**Table 1.**
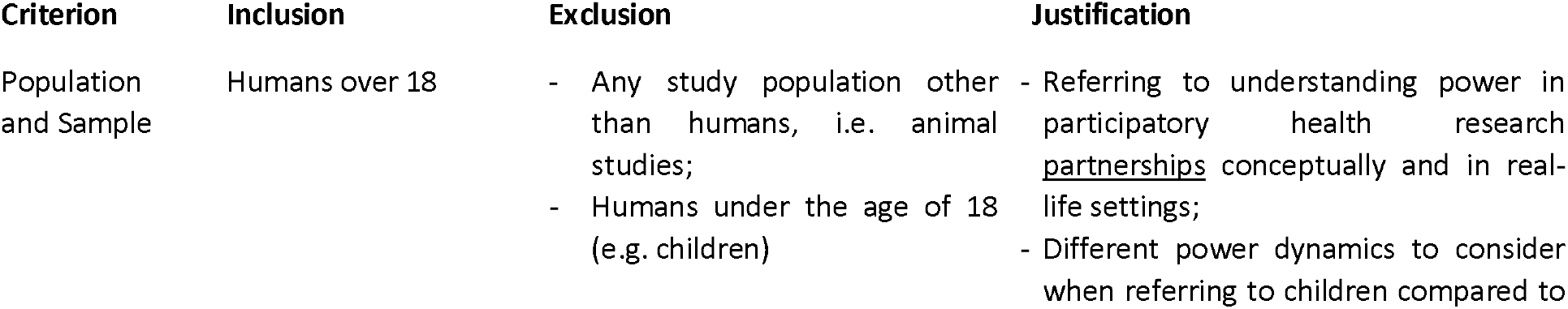
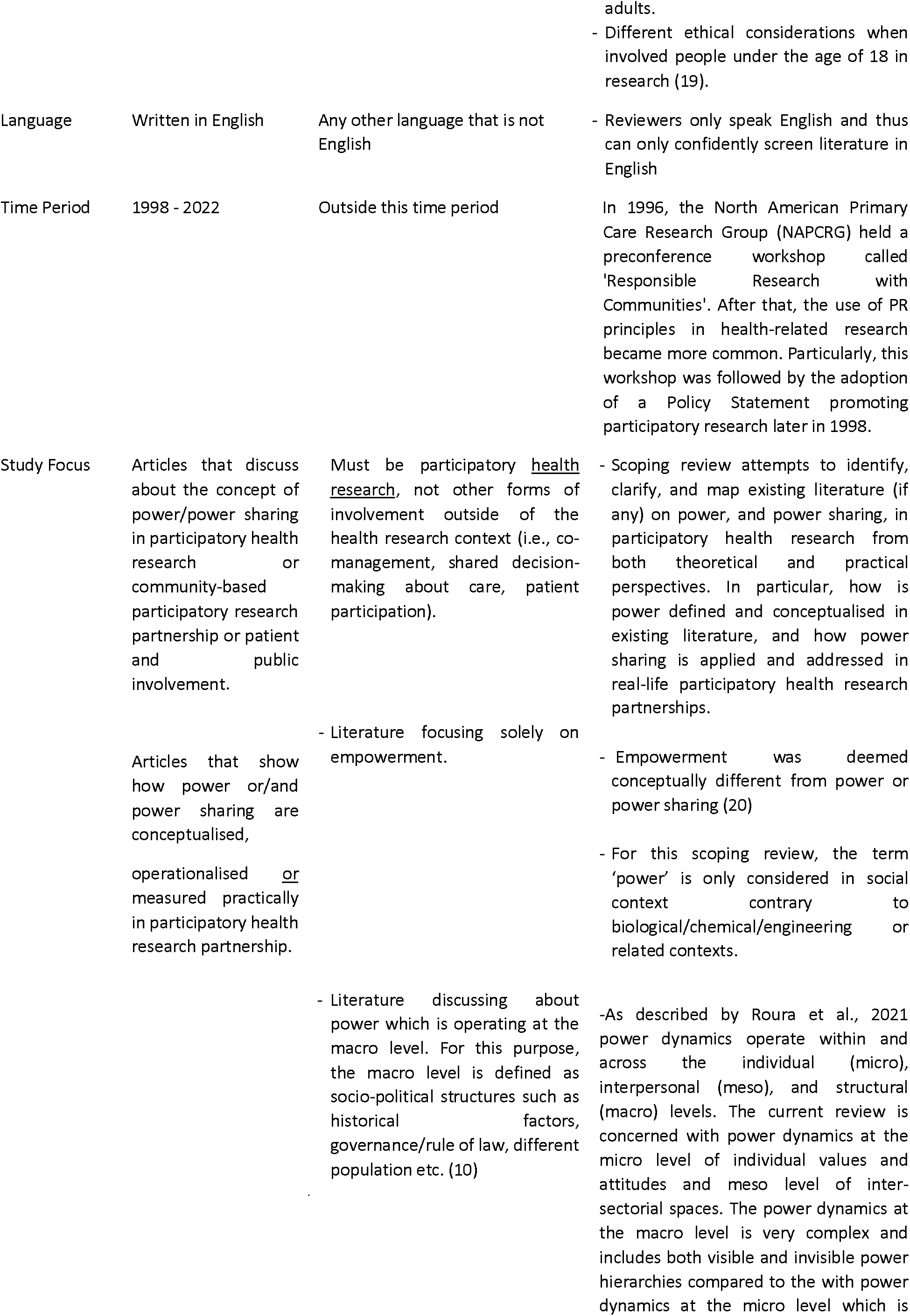
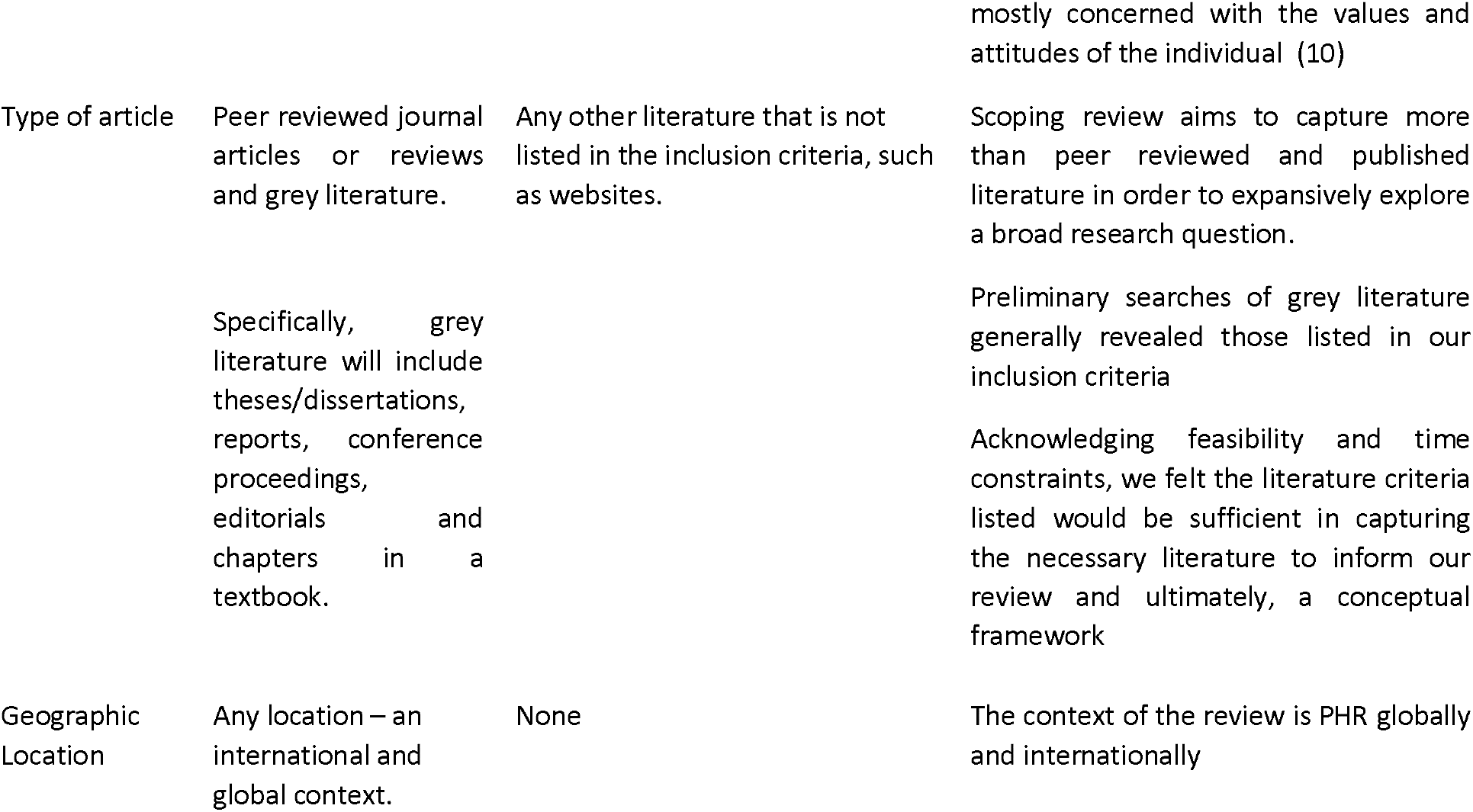
Eligibility criteria.

**Table 2.**
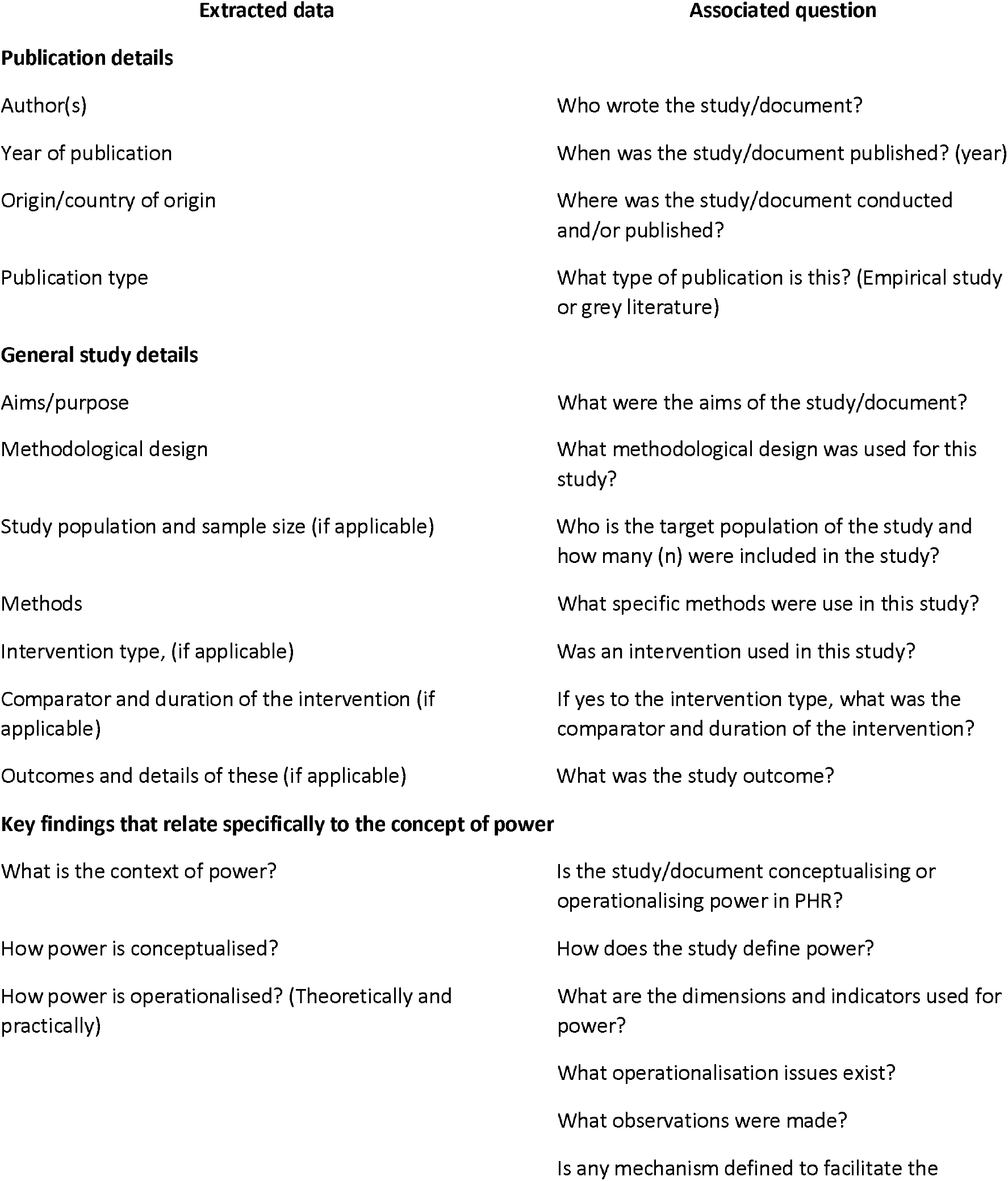
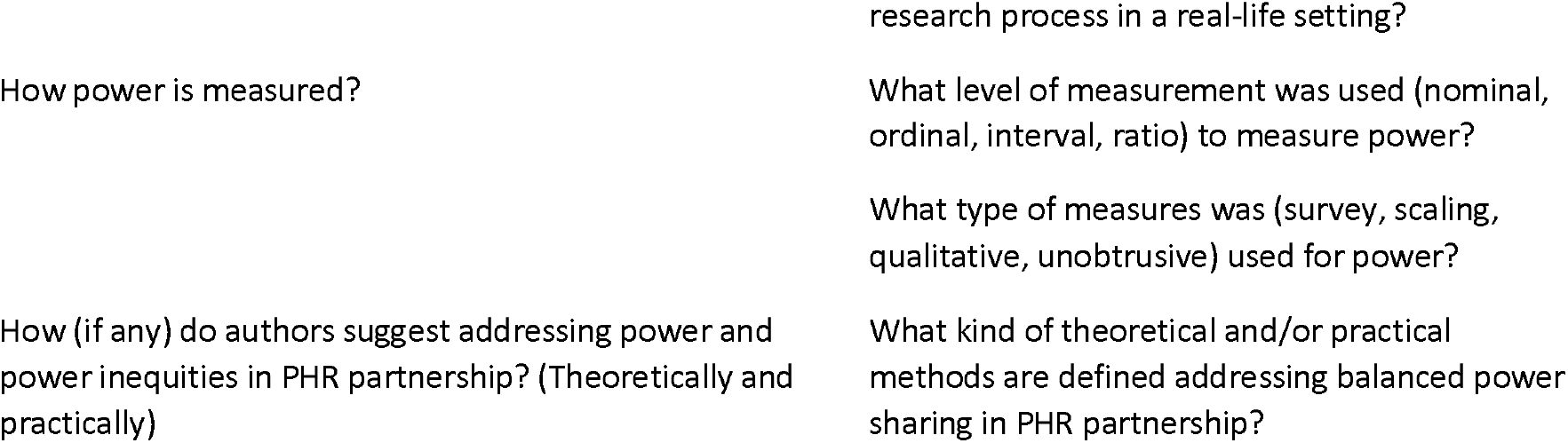
Preliminary table for data extraction and associated questions.

### Data analysis and presentation

The extracted studies will then be analysed using frequencies and counts, providing a narrative summary of the findings based on our research objectives. Our findings will be discussed collaboratively by a team of knowledge users and experts in relation to gaps in the literature and future implications. The findings will contribute to the development of a framework for assessing the level of power sharing attained in PHR projects and practical guidelines.

### Consultation with Knowledge Users

In the advanced and extended methodology of the scoping review (25), an additional stage of expert consultation (e.g. with knowledge users or experts by experience) was suggested. The present scoping review will be conducted in collaboration with two institutions (the University of Limerick and the University of Maryland, Baltimore) and a community partner providing expert knowledge by experience, (co-author BJ) residing in Baltimore, Maryland, USA. The rationale for this scoping review was derived from a previous workshop by Egid et al., (discussed above), capturing the experiences of participatory health researchers in relation to power. Findings from the workshop inspired future research in this field to gain a better understanding of the concept of power and power sharing in PHR partnerships. Afterwards, these insights will be discussed with the group of researchers from the University of Maryland, Baltimore and the community partners from the surrounding community. The detailed format for structured stakeholder discussion will be considered later. We anticipate that gaining the perspectives from non-academic stakeholders will contribute to a more complete understanding of power inequities and the co-production of knowledge in PHR.

## Data Availability

Deidentified research data will be made publicly available when the study is completed and published.

## Supporting information

S1 Appendix I. Search Strategy.

S2 Appendix II. PRISMA Flow Chart 2020.

## Author Contributions

Conceptualisation: Nino Burduladze, Laundette P. Jones, Brian D. Jones, Jon Salsberg, Meghan Gilfoyle.

Methodology: Nino Burduladze, Uchizi Msowoya, Anna Whitney.

Supervision: Meghan Gilfoyle, Jon Salsberg.

Writing – original draft: Nino Burduladze.

Writing – review & editing: Nino Burduladze, Laundette P. Jones, Brian D. Jones, Jon Salsberg, Meghan Gilfoyle, Uchizi Msowoya, Anna Whitney.

All authors have made substantive intellectual contributions to the development of this scoping review and have approved the final protocol.

## Acknowledgments

All aspects of the search strategy, including choosing the appropriate databases, was developed and conducted by Gail Betz, research and education librarian from health sciences and human services library of the University of Maryland.

## Appendix I: Search Strategy

Search strategy for PubMed conducted on 05-APR-2022

**Filters:** from 1998-present, Language - English

**Results:** 1752 studies were retrieved.

((((CBPR[Title/Abstract] OR action research[Title/Abstract] OR participative research[Title/Abstract] OR participatory research[Title/Abstract] OR co-production[Title/Abstract] OR co-researcher* OR research partnership[Title/Abstract] OR emancipator research[Title/Abstract] OR participatory rural[Title/Abstract] OR collaborative inquiry[Title/Abstract] OR decolonizing methodolog*[Title/Abstract] OR appreciative inquiry[Title/Abstract] OR dialectic* inquiry[Title/Abstract] OR cooperative inquiry[Title/Abstract] OR community-partnered action research[Title/Abstract] OR community-driven action research[Title/Abstract] OR community-driven research[Title/Abstract] OR participatory evaluation[Title/Abstract] OR community engaged research[Title/Abstract] OR community-university[Title/Abstract] OR community-academic[Title/Abstract] OR collective knowledge[Title/Abstract]) OR (“Consumer Advocacy”[Mesh] AND research)) OR (“Community-Institutional Relations”[Mesh])) OR (“Community-Based Participatory Research”[Mesh])) AND (“Power, Psychological”[Mesh] OR (“power”[Title/Abstract] OR “resource sharing”[Title/Abstract] OR “empowerment”[Title/Abstract] OR “resource allocation”[Title/Abstract] OR “shared decision making”[Title/Abstract] OR “social capital”[Title/Abstract] OR “epistemic injustice”[Title/Abstract] OR “silencing”[Title/Abstract]))

## Appendix II PRISMA Flow Chart 2020

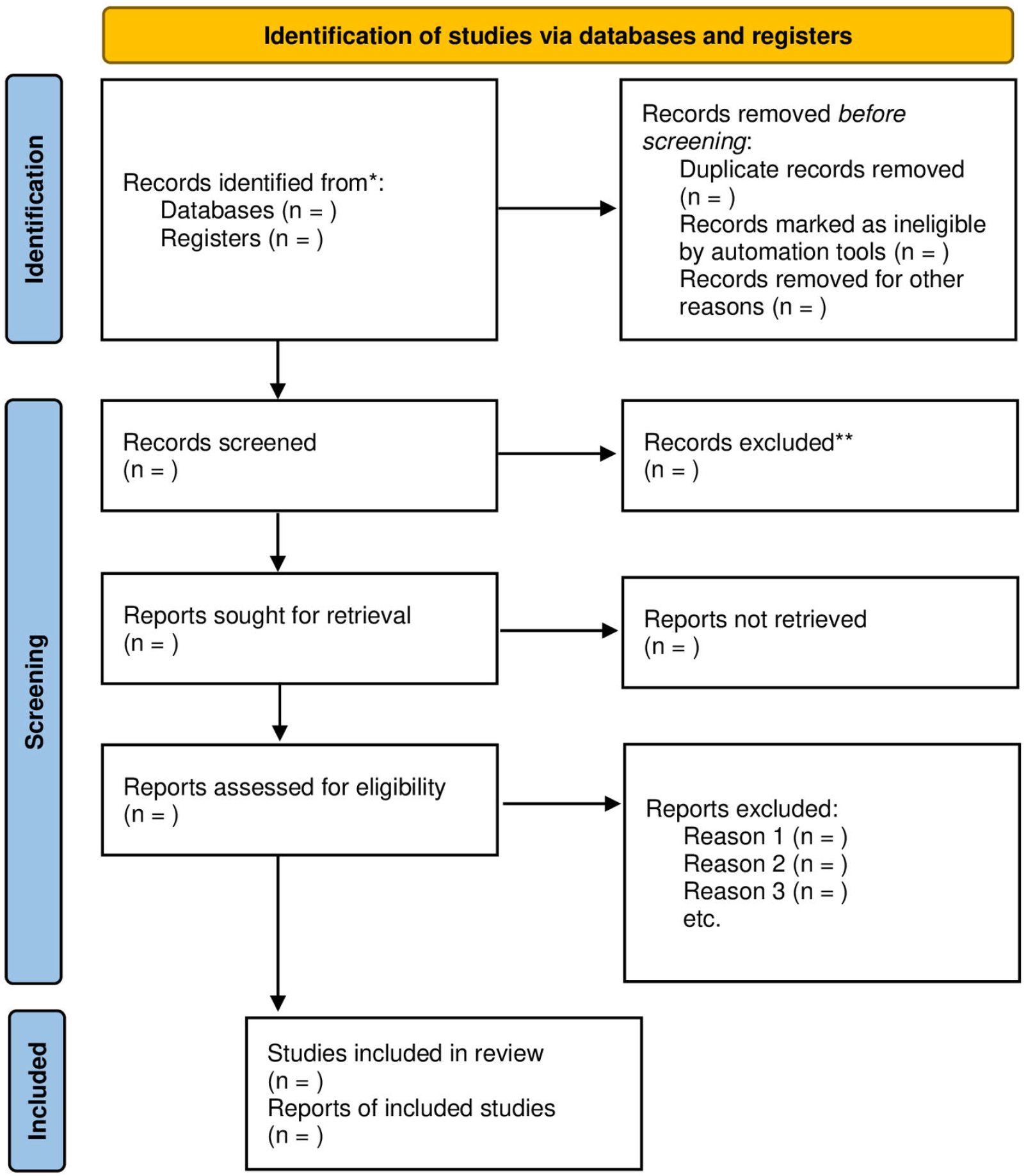

*From:* Page MJ, McKenzie JE, Bossuyt PM, Boutron I, Hoffmann TC, Mulrow CD, et al. The PRISMA 2020 statement: an updated guideline for reporting systematic reviews. BMJ 2021;372:n71. doi: 10.1136/bmj.n71

For more information, visit: https://prisma-statement.Org//

